# A Comparative Study on Deep Convolutional Neural Networks and Histogram Equalization Techniques for Glaucoma Detection From Fundus Images

**DOI:** 10.1101/2024.10.25.24316109

**Authors:** Ashish Kulkarni, H Shafeeq Ahmed

## Abstract

**Aims:** To evaluate the performance of eight different convolutional neural network (CNN) models and histogram equalization techniques for glaucoma detection from fundus images.

**Material and Methods:** The study utilized the ACRIMA database, comprising 705 fundus images (396 glaucomatous and 309 normal). The CNN architectures evaluated include VGG-16, VGG-19, ResNet-50, ResNet-152, Inception v3, Xception, DenseNet-121, and EfficientNetB7. Two histogram-based preprocessing methods were applied: histogram equalization (HE) and contrast limited adaptive histogram equalization (CLAHE). The models were trained using supervised learning, with image augmentation techniques applied. Performance metrics such as accuracy, sensitivity, specificity, precision, negative predictive value, Dice Similarity Coefficient, and Area Under the Receiver Operating Characteristics Curve (AUC ROC) were used for evaluation.

**Results:** VGG-19 achieved the highest accuracy (97.9%) in the raw data setting, followed closely by VGG-16 (97.2%). ResNet-50 and ResNet-152 showed the highest specificity scores (98.4%). The sensitivity of the models ranged from 91.3% to 98.8%, with VGG-16 and VGG-19 demonstrating the highest values. CLAHE preprocessing resulted in improved performance, particularly for ResNet-152, which achieved an accuracy of 97.5% and sensitivity of 97.9%. The AUC ROC values varied from 0.937 to 0.996, with VGG-16 having the highest value (0.998).

**Conclusion:** The study highlights the importance of selecting suitable CNN architectures and preprocessing techniques in developing effective glaucoma detection systems. While VGG-19 exhibited the highest accuracy with raw data, VGG-16 and ResNet-50 offered consistent performance across different preprocessing techniques, making them reliable options for clinical applications.

## 1 Introduction

Glaucoma is a leading cause of irreversible blindness in both the elderly and adult populace with a projected 112 million cases by 2040 [1]. Chronic neuropathy further leads to irreversible optic disk damage with structural nerve fibre damage, ultimately leading to vision loss. Characteristic changes to the optic nerve head (ONH) or the optic disk is characteristic of glaucoma [2]. The ONH is the location where ganglion cell axons form the optic disk (OD) while leaving the eye. The optic disk is visually separated into a bright and central zone in a fundus image. These are called the optic cup along with a peripheral portion called neuro-retinal rim [3]. The OD and optic cup are found in all individuals but abnormal variations in thesis of the cup relative to the OD is characteristic of glaucomatous eyes. Due to this, several different approaches have been discovered for OD and optic cup segmentation from colour fundus images [4, 5].

A convolutional neural network (CNN) is a network of self-optimizing neurons which learn directly from data [6]. CNNs are primarily used in pattern recognition within images. The current paper uses supervised learning to train the CNN model. Supervised learning is the process by which a network learns through a set of pre-labelled inputs. The parameters are updated iteratively with the goal of minimizing the model’s classification error. The usage of CNNs in image analysis can be traced back to the 1990s [7, 8]. However, the usage of CNNs in the mainstream was low until 2012, when it saw a significant increase [9]. This rise is attributed to the efficient use of GPUs, the adoption of the ReLU activation function, the usage of dropout layers and the usage of data augmentation techniques [10, 11].

The current paper discusses the performance of histogram equalization and contrast limited adaptive histogram equalization (CLAHE). They involve applying transformations on the image such that its histogram acquires a more uniform distribution, thus improving image contrast [12]. This contrast enhancement technique generally increases global contrast in the image. It can be useful when applied on dark or bright images [13]. CLAHE is a form of adaptive histogram equalization where upon performing the enhancement, the maximum value is used to clip the histogram and redistribute the intensity [14]. The current study evaluates eight different convolutional neural network (CNN) models and histogram equalization techniques for the classification of fundus images of glaucoma patients versus healthy.

## 2 Materials and Methods

### 2.1 Dataset

The images in the current study come from the ACRIMA database created by the Ministerio de Economía y Competitividad of Spain which develops automated algorithms for retinal disease assessment [15]. The database is composed of 705 fundus images (396 glaucomatous patients and 309 normal patients). All images were obtained from glaucomatous and normal patients. The present study does not require institutional ethics approval as no patients were involved in this study.

All the patients in the database were selected by a team of experts based on an inclusion and exclusion criteria formed by clinical findings on examination. Most fundus images were taken either from a previously dilated left or right eye and centred in the OD. Images were taken with a 35° field of view using the Topcon TRC retinal camera and IMAGEnet® capture System.

All fundus images from the database were separately annotated by two glaucoma experts each with eight years of experience. No other clinical information was considered while giving labels for the fundus images.

### 2.2 Network Architecture

The current paper evaluates the performance of models based on six well-known CNN architectures:

#### VGG

Introduced in 2014, VGG was a novel approach to increasing network depth by using very small convolution filters (3 * 3) to between 16 and 19 layers. The current paper utilizes the VGG-16 and VGG-19 networks, which are of depth 16 and 19 layers respectively. [15]

#### ResNet

The ResNet architecture is a CNN that was introduced in 2015 by Microsoft Research which made use of residual learning framework to more efficiently train deeper networks. Residual nets, while much deeper than VGG nets, are of less complexity. The current paper makes use of two residual nets: ResNet-50, which has a depth of 50 layers and ResNet-152, which has a depth of 152 layers. [17]

#### Inception-v3

Inception v3 is the third edition of Google’s Inception CNN used in image analysis and object detection, introduced as a module for GoogLeNet. It is a 42-layer CNN with 11 Inception modules. In essence, the Inception module allows us to use multiple filter sizes in each image block. [18]

#### Xception

Xception, or “extreme Inception”, is a 71-layer CNN which uses depthwise separable convolutions rather than standard Inception modules. [19]

#### DenseNet

The Dense Convolutional Network, or DenseNet architecture, connects each layer to every other layer in a feed-forward fashion. The current paper uses a DenseNet with a depth of 121 layers. [20]

#### EfficientNet

EfficientNet is an architecture that leverages a different scaling method to scale all dimensions using a compound coefficient. In this study, we make use of EfficientNet-B7, which consists of approximately 66M parameters. [21]

Every architecture used is transferred with their ImageNet weights, and only the last 3 convolutional layers are set to trainable. In addition, we added two dense layers and a dropout layer in between, with a rate of 0.5.

### 2.3 Data Preprocessing

The current paper has evaluated two methods of histogram-based image preprocessing: histogram equalisation and CLAHE.

#### Histogram Equalization

This contrast enhancement technique generally increases global contrast in the image. It can be useful when applied on dark or bright images [22].

#### Contrast Limited Adaptive Histogram Equalization

CLAHE is a form of adaptive histogram equalization where upon performing the enhancement, the maximum value is used to clip the histogram and redistribute the intensity [23]. In case of performing CLAHE on a true-colour image, the CIELAB colour space is used.

### 2.4 Model Training

Each model was fine-tuned on the training date for 128 epochs, with a batch size of 32. Before feeding the training images into the neural network, image augmentation by random affine transformations (flipping, rotation and translation) and contrast adjustment is performed. Figure 2 shows the various alterations of the image after augmentation has been applied.

**Figure 1:**
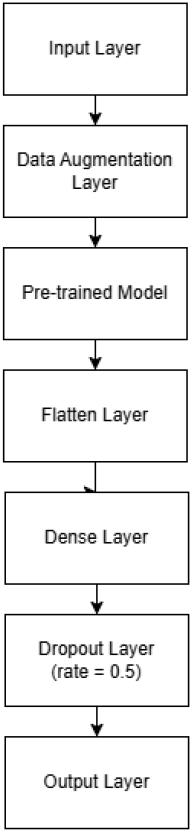
CNN architecture used.

**Figure 2:**
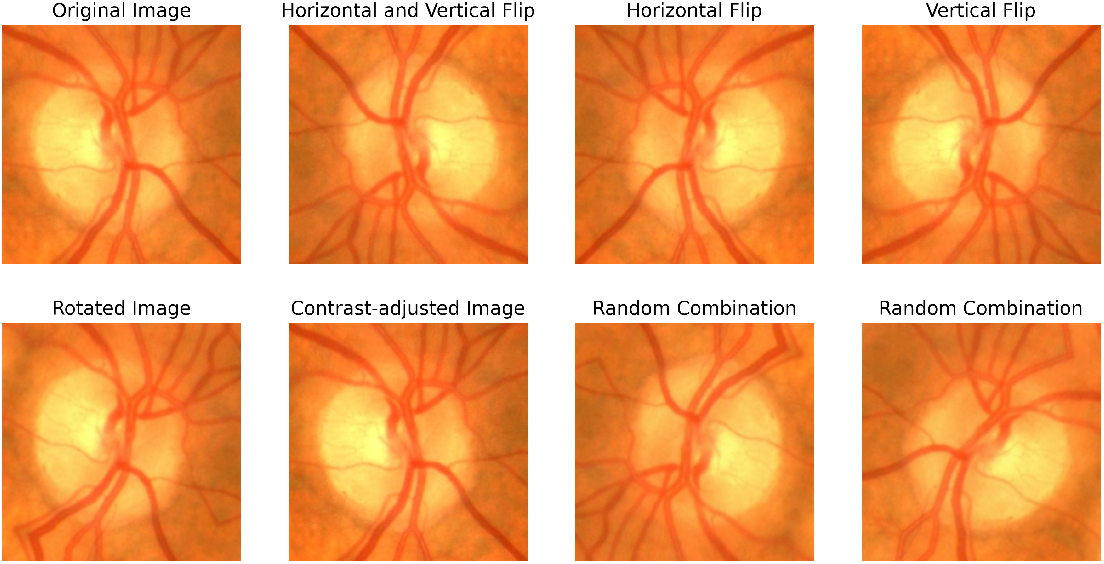
A fundus image with different transformations applied to it.

### 2.5 Model Evaluation

The present study uses several performance metrics to evaluate the models. These metrics include accuracy, which measures the overall correctness of the model’s predictions, and sensitivity (True Positive Rate), which assesses the model’s ability to correctly identify positive cases. Specificity (True Negative Rate) evaluates the model’s capability to correctly identify negative cases, while precision (Positive Predictive Value) measures the accuracy of positive predictions. Negative Predictive Value assesses the accuracy of negative predictions. The study also utilizes the Dice Similarity Coefficient (F1 Score) to provide a balance between precision and sensitivity. Finally, the Area Under the Receiver Operating Characteristics Curve (AUC ROC) is used to measure the model’s ability to distinguish between positive and negative cases across different threshold settings.

## 3 Results

Table 1 presents the performance metrics of various models without any histogram equalisation. The models under consideration are VGG-16, VGG-19, ResNet-50, ResNet-152, Inception v3, Xception, DenseNet-121, and EfficientNetB7. The accuracy values range from 0.908 to 0.979. VGG-19 achieved the highest accuracy (0.979), closely followed by VGG-16 (0.972). Specificity values range from 0.887 to 0.984. ResNet-50 and ResNet-152 have the highest specificity scores (0.984). Sensitivity, ranges from 0.913 to 0.988. VGG-16 and VGG-19 both demonstrate the highest sensitivity values (0.988). VGG-19 leads with the highest F1 score (0.981). The AUC-ROC values vary from 0.937 to 0.996. VGG-16 has the highest AUC-ROC value (0.998). The table also presents a comparison of model accuracies with the number of parameters used in each model. Among the models, VGG-19 achieved the highest accuracy of 97.89% with 28,413,762 parameters, while EfficientNetB7 had the most parameters (106,041,497) but an accuracy of 92.25%. ResNet-50 and DenseNet-121 achieved an accuracy of 95.77% and 95.77%, respectively, with significantly fewer parameters (57,142,914 and 23,815,490).

**Table 1:** Performance Metrics of Convolutional Neural Network Models without Histogram Equalization.

| Model | Accuracy | Specificity | Sensitivity | F1 Score | Area Under ROC | Number of Parameters |
| --- | --- | --- | --- | --- | --- | --- |
| *VGG-16* | 0.9718 | 0.9516 | 0.9875 | 0.9753 | 0.9978 | 2,31,04,066 |
| *VGG-19* | 0.9789 | 0.9677 | 0.9875 | 0.9814 | 0.9933 | 2,84,13,762 |
| *ResNet-50* | 0.9577 | 0.9839 | 0.9375 | 0.9615 | 0.9956 | 5,71,42,914 |
| *ResNet-152* | 0.9507 | 0.9839 | 0.925 | 0.9548 | 0.9944 | 9,19,26,146 |
| *Inception v3* | 0.9085 | 0.8871 | 0.925 | 0.9193 | 0.9364 | 4,06,77,922 |
| *Xception* | 0.9296 | 0.9516 | 0.9125 | 0.9359 | 0.9794 | 5,44,16,682 |
| *DenseNet-121* | 0.9577 | 0.9516 | 0.9625 | 0.9625 | 0.9960 | 2,38,15,490 |
| *EfficientNetB7* | 0.9225 | 0.8871 | 0.95 | 0.9325 | 0.9625 | 10,60,41,497 |

Table 2 displays performance metrics of CNNs with histogram equalization (HE). The VGG-16 model demonstrated high accuracy and sensitivity. Its performance was closely matched by ResNet-50, which also showed high accuracy and a slightly lower sensitivity but maintained a high specificity and AUC. VGG-19, while having good specificity, showed lower sensitivity compared to VGG-16 and ResNet-50. ResNet-152 exhibited a balanced performance with high accuracy, sensitivity, and specificity. Inception v3 and Xception, although less accurate compared to the others, still performed well, with Inception v3 showing relatively lower sensitivity. DenseNet-121 and EfficientNetB7 also performed well, with DenseNet-121 achieving high sensitivity and EfficientNetB7 balancing between accuracy and the number of parameters.

**Table 2:**
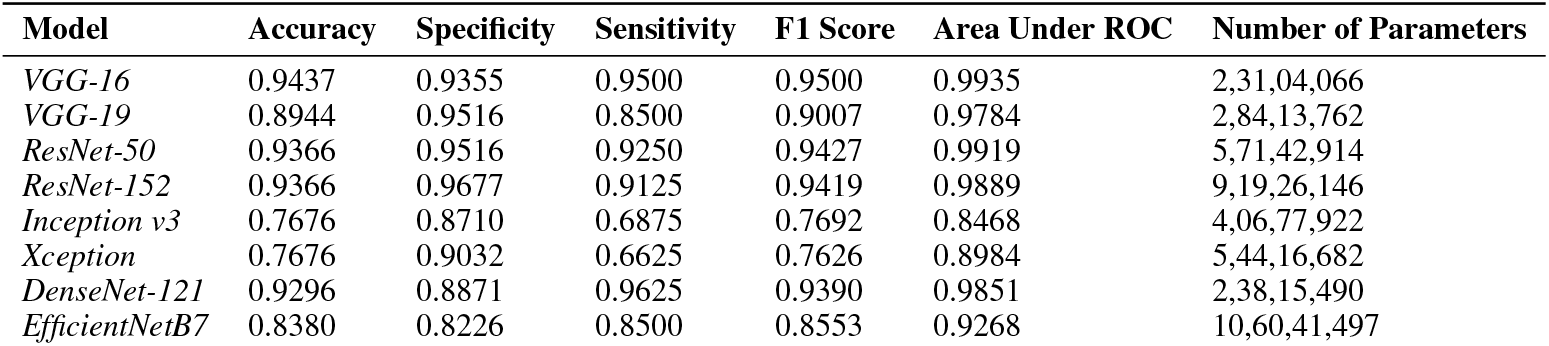
Performance Metrics of Convolutional Neural Network Models with Histogram Equalization.

Table 3 displays performance metrics of CNNs with CLAHE. ResNet-152 achieved the highest accuracy and sensitivity, significantly improving upon its histogram equalization results. VGG-16 and VGG-19 both maintained their performance with perfect specificity but had slightly lower sensitivity compared to their HE results. ResNet-50 also showed improved sensitivity with CLAHE. DenseNet-121 stood out with high accuracy and sensitivity, rivalling the performance of ResNet-152. EfficientNetB7 and Xception showed moderate improvements, while Inception v3 had a balanced yet slightly lower performance compared to the other models.

**Table 3:** Performance Metrics of Convolutional Neural Network Models with CLAHE.

| Model | Accuracy | Specificity | Sensitivity | F1 Score | Area Under ROC | Number of Parameters |
| --- | --- | --- | --- | --- | --- | --- |
| *VGG-16* | 0.9225 | 1.0000 | 0.8625 | 0.9262 | 0.9966 | 2,31,04,066 |
| *VGG-19* | 0.9225 | 1.0000 | 0.8625 | 0.9262 | 0.9966 | 2,84,13,762 |
| *ResNet-50* | 0.9296 | 0.8710 | 0.9750 | 0.9398 | 0.9838 | 5,71,42,914 |
| *ResNet-152* | 0.9577 | 0.9355 | 0.9750 | 0.9630 | 0.9942 | 9,19,26,146 |
| *Inception v3* | 0.8099 | 0.7742 | 0.8375 | 0.8323 | 0.9173 | 4,06,77,922 |
| *Xception* | 0.7817 | 0.9355 | 0.6625 | 0.7737 | 0.9212 | 5,44,16,682 |
| *DenseNet-121* | 0.9507 | 0.9355 | 0.9625 | 0.9565 | 0.9877 | 2,38,15,490 |
| *EfficientNetB7* | 0.8944 | 0.8387 | 0.9375 | 0.9091 | 0.9256 | 10,60,41,497 |

Figure 3 compares the average values of the models across different preprocessing techniques. The Normal preprocessing technique yielded the highest average performance with an accuracy of 94.01%, sensitivity of 95.63%, F1 score of 94.75%, and AUC of 97.92%. Histogram Equalization showed slightly lower performance, with an accuracy of 88.38%, sensitivity of 90.63%, F1 score of 89.72%, and AUC of 95.59%. CLAHE offered a balanced performance, with an accuracy of 92.25%, sensitivity of 95.00%, F1 score of 93.28%, and AUC of 95.67%, outperforming HE but slightly behind Normal preprocessing.

**Figure 3:**
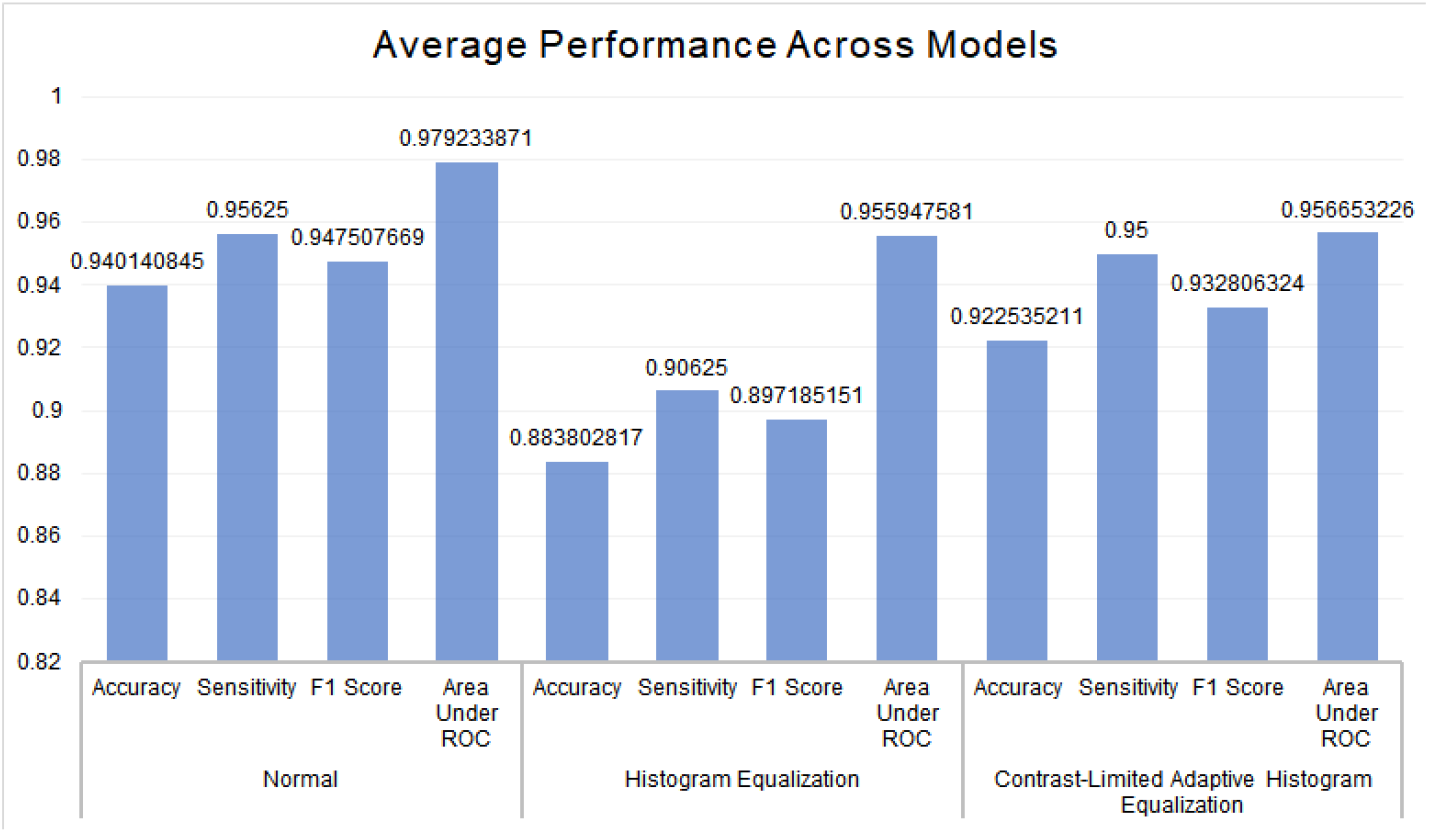
Accuracy of classification across different preprocessing techniques.

Figure 4 illustrates the ROC curves for various deep learning models used in the study. The VGG-16, ResNet50, and DenseNet-121 models achieved an AUC of 1.00. VGG-19 and ResNet-152 closely followed with AUCs of 0.99. The other models while having lower AUCs, also performed well.

**Figure 4:**
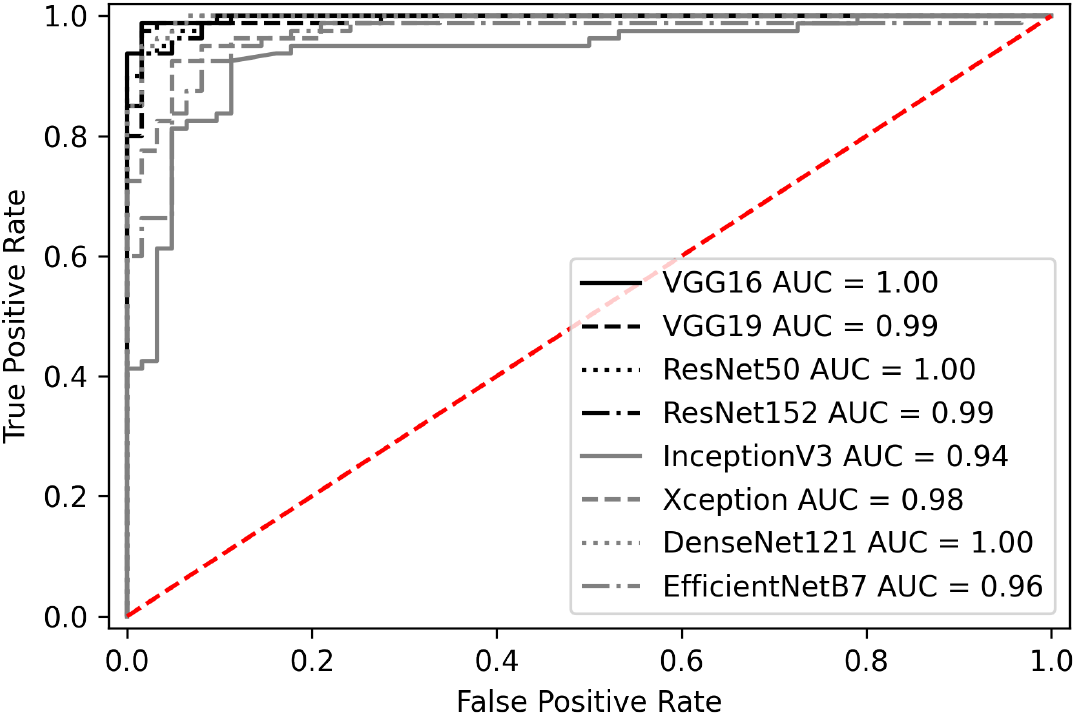
Receiver operating characteristic curves of each classifier.

## 4 Discussion

The current study aimed to evaluate the performance of an AI model for glaucoma classification using convolutional neural networks (CNNs). The results demonstrated that CNN models, particularly VGG-19 and VGG-16, showed the highest accuracies and sensitivities in distinguishing between glaucoma patients and healthy individuals, outperforming other architectures like ResNet, Inception v3, Xception, DenseNet-121, and EfficientNetB7. These findings are significant as they indicate the potential of AI-based systems in assisting with the early detection and diagnosis of glaucoma, which is a leading cause of irreversible blindness. The high accuracy achieved by the deep learning model suggests its capability to correctly identify positive cases, thereby aiding in timely interventions and improved patient outcomes. To the best of our knowledge this is the first study using HE and CLAHE in glaucoma evaluation.

Among the CNN models evaluated, VGG-19 emerged with the highest accuracy in the raw data setting, which has been a major model used by other researchers. As seen in our study, coloured fundus photographs happen to be the preferred option [24, 25, 26]. However, its performance declined notably when HE was applied, probably due to its sensitivity to preprocessing methods. Conversely, models like VGG-16 and ResNet-50 demonstrated more stable performance across different preprocessing techniques. However, several author when comparing HE and CLAHE with other techniques have also not noted a significant improvement [27]. On the other hand Alshamrani et al. [28] were able to get beneficial outcomes from the use of histogram equalisation techniques for three CNN classification.

HE and CLAHE techniques were employed to enhance image quality and improve model performance. CLAHE works by removing noise and preventing noise amplification as well [29, 30]. While HE initially showed promise in enhancing image contrast, our results indicate mixed outcomes across different CNN models. For instance, CLAHE demonstrated a more consistent improvement in sensitivity metrics compared to HE, reiterating its potential in enhancing image features critical for glaucoma detection. However, the methodology behind its working is easily dependent on the images themselves rather than the algorithm [31, 32].

The evaluation of model size relative to performance metrics showed interesting trade-offs. Despite the higher parameter counts of models like EfficientNetB7, their performance did not consistently outpace that of smaller architectures like VGG-16 and ResNet-50. This suggests that while model complexity is important, architectural design and sensitivity to preprocessing techniques play critical roles in achieving high accuracy and reliability. Additionally, the number of parameters in a model, or the model’s size, often plays a large role in the deployability of the solution [33]. A large model would be harder to run onsite, and would take longer to process an input [34]. However, it is important to note that regardless of the methodology followed or issues with optimisation, our model performed superior to the majority of the CNN models evaluated insofar for glaucoma detection from colour fundus images [35].

The present study does have its limitations, including the utilisation of a third-party database. Moreover, the images were clearly cropped with a focus on the optic disc, which is often not seen in clinical practice as these can be obtained from various angles. However, the findings of this study have important implications for clinical practice. By identifying CNN models and preprocessing techniques that optimize sensitivity and specificity for glaucoma detection, clinicians can potentially enhance diagnostic accuracy and efficiency. Future research directions may explore hybrid preprocessing methods or novel architectures tailored to specific image characteristics inherent in fundus images, further refining the capabilities of automated glaucoma detection systems.

## 5 Conclusion

In conclusion, our comparative study shows the importance of selecting appropriate CNN architectures and preprocessing techniques in the development of robust glaucoma detection systems. While VGG-19 initially demonstrated superior accuracy, its sensitivity to preprocessing methods calls for careful consideration in clinical applications. Models like VGG-16 and ResNet-50, with their balanced performance across different settings, offer practical solutions for reliable glaucoma detection. Moving forward, continued advancements in CNN design and image enhancement techniques hold promise for further improving diagnostic outcomes in ophthalmology.

## Data Availability

All data produced in the present study are available upon reasonable request to the authors
All data produced in the present work are contained in the manuscript

